# Quantity of SARS-CoV-2 RNA copies exhaled per minute during natural breathing over the course of COVID-19 infection

**DOI:** 10.1101/2023.09.06.23295138

**Authors:** Gregory Lane, Guangyu Zhou, Judd F. Hultquist, Lacy M. Simons, Ramon Lorenzo- Redondo, Egon A. Ozer, Danielle M. McCarthy, Michael G. Ison, Chad J. Achenbach, Xinkun Wang, Ching Man Wai, Eugene Wyatt, Alan Aalsburg, Qiaohan Yang, Torben Noto, Arghavan Alisoltani, Daniel Ysselstein, Rajeshwar Awatramani, Robert Murphy, Grant Theron, Christina Zelano

## Abstract

SARS-CoV-2 is spread through exhaled breath of infected individuals. A fundamental question in understanding transmission of SARS-CoV-2 is how much virus an individual is exhaling into the environment while they breathe, over the course of their infection. Research on viral load dynamics during COVID-19 infection has focused on internal swab specimens, which provide a measure of viral loads inside the respiratory tract, but not on breath. Therefore, the dynamics of viral shedding on exhaled breath over the course of infection are poorly understood. Here, we collected exhaled breath specimens from COVID-19 patients and used RTq-PCR to show that numbers of exhaled SARS-CoV-2 RNA copies during COVID-19 infection do not decrease significantly until day 8 from symptom-onset. COVID-19-positive participants exhaled an average of 80 SARS-CoV-2 viral RNA copies per minute during the first 8 days of infection, with significant variability both between and within individuals, including spikes over 800 copies a minute in some patients. After day 8, there was a steep drop to levels nearing the limit of detection, persisting for up to 20 days. We further found that levels of exhaled viral RNA increased with self-rated symptom-severity, though individual variation was high. Levels of exhaled viral RNA did not differ across age, sex, time of day, vaccination status or viral variant. Our data provide a fine-grained, direct measure of the number of SARS-CoV-2 viral copies exhaled per minute during natural breathing—including 312 breath specimens collected multiple times daily over the course of infection—in order to fill an important gap in our understanding of the time course of exhaled viral loads in COVID-19.

## INTRODUCTION

SARS-CoV-2, the causative agent of COVID-19, spreads through exhaled breath during coughing, talking, singing, and breathing (1–7). Levels of SARS-CoV-2 over the course of infection have been extensively characterized in upper and lower respiratory tract specimens such as nasopharyngeal and oropharyngeal swabs (8–11, 38–39), whereas the dynamics of levels on breath over the course of infection remain virtually unexplored. This is despite the fact that quantifying levels of viral shedding on exhaled breath would allow for a direct approximation of the amount of virus an individual is shedding into the environment (12–17), thereby exposing others to risk of infection. We know particularly little about the dynamics of viral shedding on breath during unlabored natural breathing, which serves as a baseline for viral transmission on breath.

While the dynamics of viral load inside the host respiratory tract has direct relevance to viral pathology, dynamics of viral load on the host’s breath has direct relevance to infectiousness. Understanding the dynamics of viral shedding on breath is important for prevention of transmission of disease. Measuring viral load characteristics of the primary route of onward transmission is critical to inform isolation times in the clinic, where isolation consumes scarce resources, and to inform public health transmission control protocols. In addition, variables that impact level of viral shedding on breath remain unclear, but may vary by a multitude of factors including symptom severity, days since symptom onset, co-morbidities, viral genotype, and other unknowns. Understanding of these factors requires quantification of exhaled viral loads, which cannot be inferred from internal viral loads.

Current techniques for measuring viral load in exhaled breath have successfully detected SARS-CoV-2 in specimens (18–27), though with variable detection rates ranging from 26.9% to 86%. Recent work has also shown that SARS-CoV-2 can be isolated from exhaled breath, confirming that it contains replication-competent virus (17). However, prior work has focused on exertive breathing conditions (talking, singing, coughing), and we therefore have less understanding of viral loads in exhaled breath during natural breathing. Furthermore, prior work has not analyzed exhaled breath collected longitudinally over the course of infection relative to the day of symptom onset (potentially due to expense and lack of portability of breath collection devices), which would allow for a better understanding of the time course of changes in viral loads on breath. An inexpensive, portable device that allows patients to self-collect breath samples at home would facilitate research into factors that contribute to virus transmission from breath, how exhaled virus levels change over the course of infection, and whether therapeutics and other interventions reduce levels of exhaled virus.

Here, we developed a portable, disposable exhaled breath condensate collection device (EBCD) (**Fig S1**) and used it to collect 312 specimens from 60 COVID-19-tested outpatients who were treated at Northwestern Memorial Hospital (NMH). Specimens were analyzed using RT-qPCR. Our data set included breath specimens collected multiple times per day over the course of infection. We report numbers of SARS-CoV-2 RNA copies exhaled per minute, during natural breathing, over the course of infection and across a range of factors including self-reported symptom severity, age, sex, presence of co-morbidities, vaccination status and viral variant.

## RESULTS

### Detection of SARS-CoV-2 RNA in exhaled breath

To collect exhaled breath specimens, we developed a new exhaled breath collection device (EBCD) that is inexpensive, portable and disposable, allowing at-home collection of samples by participants. The device consists of a repurposed syringe tube fitted with one-way valve and mouth piece (**Fig S1**). A cooling sleeve placed over the tube causes exhaled breath to condense on the inner wall during a 10-minute breath collecting session. Participants were instructed to breathe naturally out of their mouth through the tube at a normal pace. To validate our device as a tool to measure SARS-CoV-2 in exhaled breath, we collected specimens from 60 study participants recruited after receiving a clinical, PCR-based SARS-CoV-2 diagnostic test via nasopharyngeal swab (typically Abbott ID NOW platform; see **Table 1** for patient demographics), within 10 days of symptom onset. We analyzed our specimens with RT-qPCR. SARS-CoV-2 RNA was detected in 43 of the 43 breath specimens collected from participants who were positive for COVID-19 by diagnostic testing conducted within the first 10 days from symptom onset (100%). SARS-CoV-2 RNA was not detected in any of the 16 participants who were negative for COVID-19 by diagnostic testing (100%) (**Fig 1A**). Thus, the correspondence between our test and the clinical diagnostic test was 100% accurate within the first 10 days of symptoms. The total number of SARS-CoV-2 RNA copies detected over the 10-minute breathing session ranged from 5 to 8757, with a mean of 834 (95% CI: [259, 1409]). To confirm the specificity of the amplified target, sequencing of resultant PCR product was performed on two specimens, validating proper target amplification. To confirm that the EBCD collects breath condensate without contamination from saliva, we performed viscosity measurements on EBCD samples collected during nasal breathing and oral breathing separately and compared them to viscosity measurements of saliva and water. If oral breath samples collected with EBCD were contaminated with saliva, then their viscosity should be increased compared to nasal breath samples. We found no statistical difference between the viscosity of nasal and oral breath samples, or between either sample type and water, validating our sample type as exhaled breath condensate (**Fig S2**). Thus, EBCD accurately detects SARS-CoV-2 RNA in exhaled breath.

**Fig 1.**
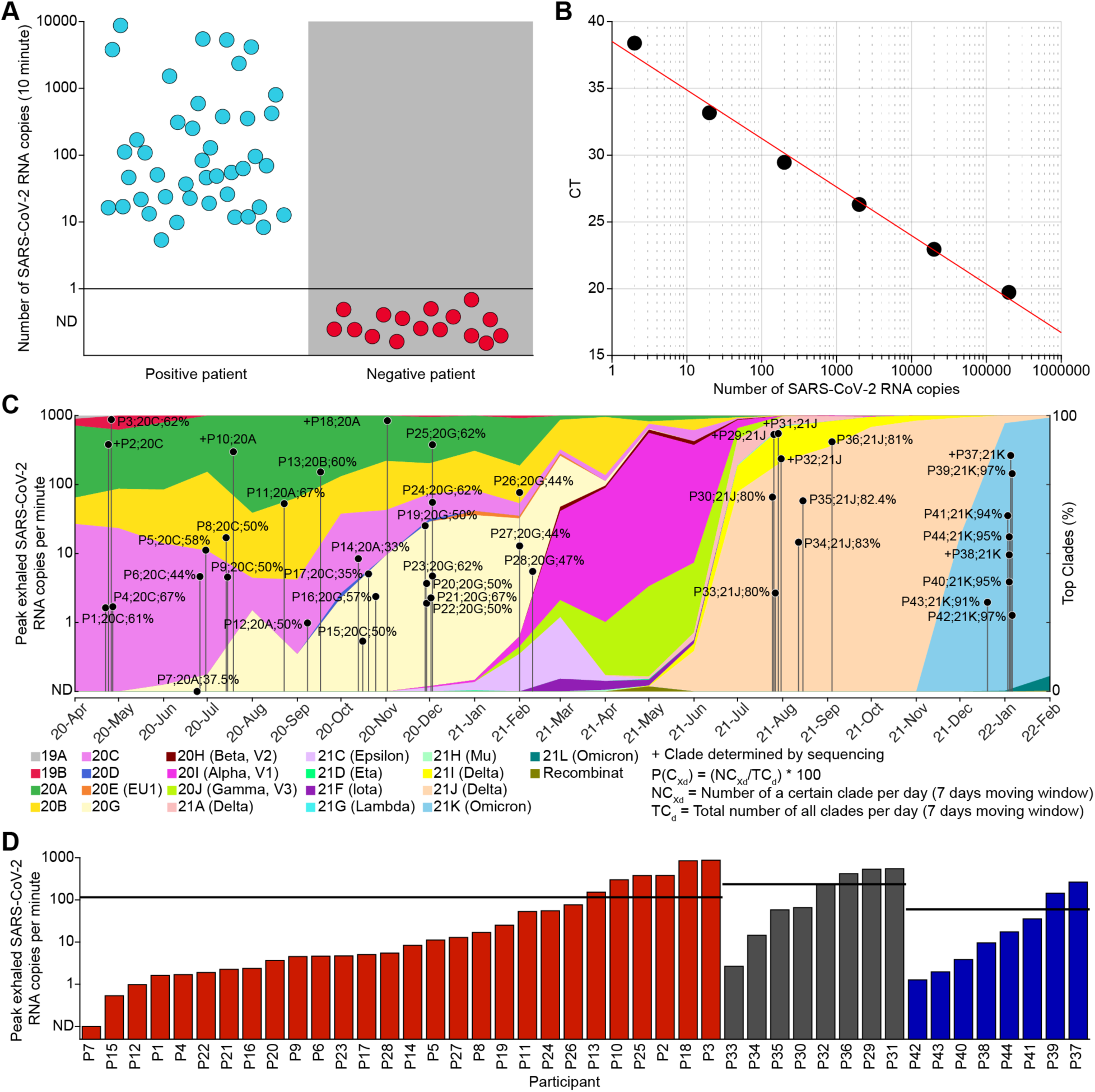
(A) Comparison of EBCD results with clinical tests for COVID-19. Results are shown for 60 patients who used the EBCD to produce a sample after they were tested for COVID-19 using nasal swab samples and the Abbott ID NOW nucleic acid-based rapid diagnostic test (n = 42), or RT-qPCR (n = 18). 44 patients tested positive on the clinical test (blue dots) and 16 patients tested negative on the clinical test (red dots). EBCD results are plotted as the number of exhaled SARS-CoV-2 RNA copies over the 10-minute breathing session. (B) Dilution series. Dilution series were used to convert cycle threshold values to viral copies. (C) Identification of SARS-CoV-2 variant by clade for participant samples over time. Background colors represent SARS-CoV-2 variant clade percentage in Chicago between April 2020 and February 2022. Dots represent peak amount of viral RNA detected on breath for each COVID-19-positive participant. Participant number, putative clade and percentage likelihood are indicated next to each dot; +, clade identified by genome sequencing; vertical lines indicate day of symptom onset (n = 42) or date of diagnosis (n = 2; P15 and P43). (D) Peaks levels of viral RNA shedding on breath across participants, separated by variant and sorted by exhaled viral RNA copy number. Black horizontal lines indicate average within each variant type. Red, pre-Delta; Gray, Delta; Blue, Omicron.

**Table 1.** COVID-19-positive participant demographics.

| Variable | Descriptor | Female | Male |
| --- | --- | --- | --- |
| Total No. | -- | 20(45.5) | 24(54.5) |
| Age | Median (IQR) | 32.0(18.0) | 37.0(16.5) |
| Race |  |  |  |
|  | White | 8(40.0) | 13(54.2) |
|  | Black or African American | 6(30.0) | 7(29.2) |
|  | Unknown | 0(0.0) | 2(8.3) |
|  | Asian | 3(15.0) | 1(4.2) |
|  | Other | 3(15.0) | 1(4.2) |
| Ethnicity |  |  |  |
|  | Not Hispanic or Latino | 17(85.0) | 21(87.5) |
|  | Hispanic or Latino | 3(15.0) | 2(8.3) |
|  | Unknown | 0(0.0) | 1(4.2) |
| Smoking |  |  |  |
|  | Never | 11(55.0) | 12(50.0) |
|  | Former | 4(20.0) | 2(8.3) |
|  | Missing | 3(15.0) | 6(25.0) |
|  | Current | 2(10.0) | 4(16.7) |
| Vaccine status at presentation |  |  |  |
|  | None | 14(70.0) | 17(70.8) |
|  | Full | 5(25.0) | 6(25.0) |
|  | Partial | 1(5.0) | 1(4.2) |
| Clinic of presentation |  |  |  |
|  | NMH ED | 13(65.0) | 18(75.0) |
|  | NMH ICCs | 5(25.0) | 6(25.0) |
|  | Community Health Clinic | 2(10.0) | 0(0.0) |
| Clinical test |  |  |  |
|  | Abbott ID NOW | 14(70.0) | 20(83.3) |
|  | RT-qPCR | 6(30.0) | 4(16.7) |
| Clinical disposition |  |  |  |
|  | Outpatient | 19(95.0) | 23(95.8) |
|  | Inpatient | 1(5.0) | 1(4.2) |
| Comorbidity at presentation |  |  |  |
|  | None | 15(75.0) | 22(91.7) |
|  | Asthma | 3(15.0) | 1(4.2) |
|  | Diabetes | 1(5.0) | 1(4.2) |
|  | Cancer | 1(5.0) | 0(0.0) |
| Complaints at presentation |  |  |  |
|  | Other | 17(85.0) | 23(95.8) |
|  | Fever | 6(30.0) | 14(58.3) |
|  | Cough | 9(45.0) | 11(45.8) |
|  | Body aches | 5(25.0) | 9(37.5) |
|  | Headache | 10(50.0) | 8(33.3) |
|  | Shortness of breath | 7(35.0) | 6(25.0) |
|  | Chest pain | 2(10.0) | 6(25.0) |
|  | Chills | 4(20.0) | 6(25.0) |
|  | Fatigue | 2(10.0) | 5(20.8) |
|  | Loss of taste/smell | 1(5.0) | 4(16.7) |
|  | Sore throat | 7(35.0) | 4(16.7) |
|  | None | 3(15.0) | 1(4.2) |
| Date of diagnosis |  |  |  |
|  | April–June 2020 | 1(5.0) | 3(12.5) |
|  | July–September 2020 | 4(20.0) | 5(20.8) |
|  | October–December 2020 | 6(30.0) | 6(25.0) |
|  | January–March 2021 | 1(5.0) | 2(8.3) |
|  | July–September 2021 | 3(15.0) | 5(20.8) |
|  | January–March 2022 | 5(25.0) | 3(12.5) |

Data are represented as No. (%), except for Age, which is represented as median (IQR, interquartile range). NMH ED, Northwestern Memorial Hospital Emergency Department; NMH ICCs, Northwestern Memorial Hospital Immediate Care Clinics.

#### Average numbers of SARS-CoV-2 RNA copies on breath did not decrease until day 8 from symptom onset

To examine the dynamics of exhaled viral loads over the course of infection, we invited COVID-19-positive individuals to participate in voluntary longitudinal home collection of additional exhaled breath specimens, typically two per day, for a duration determined by participant availability (ranging from 2 to 20 days, see Table S1 for the number of samples provided by each participant). At the time of each at-home sample collection session, participants recorded time of day, current symptoms, and subjective self-assessed overall symptom severity (resolved/none, mild, moderate, severe). This resulted in a total of 294 specimens, each associated with a day-since-symptom-onset and symptom-severity score. To examine the time-course of viral shedding on exhaled breath, we plotted SARS-CoV-2 viral copies against days since symptom onset for all samples, which showed that the number of SARS-CoV-2 viral copies exhaled per minute decreased sharply on day 8 (**Fig 2A**). To quantify this effect, we fitted the moving-average time course of viral RNA exhaled per minute to an S curve model (R^2^ = 0.7657, adjusted R^2^ = 0.7567), which yielded a modelled inflection point of maximal decrease at day 8 (RMSE: 0.3755) (**Fig 2A**, green dashed line). In a second confirmatory analysis, samples were binned according to their days from symptom onset (1–2, 3–4, 5–6, 7–8, 9–10, 11–12 days) and mean exhaled viral RNA copies were computed for each bin (bin:(mean [%95 CI]); 1–2 days: 37 [6, 68]; 3–4 days: 80 [43, 117]; 5–6 days: 58 [19, 97]; 7–8 days: 13 [4, 23]; 9–10 days: 2 [1, 2]; 11–12 days: 2 [1, 3]) (**Fig 2B**). Levels were similar across bins including days 1 to 6 (Tukey test, all P’s > 0.05), with the first significant difference observed between the third (days 5-6 from symptom onset) and fourth (days 7-8 from symptom onset) bins (Tukey test, *P* < 0.05). Taken together, these findings suggest that the number of viral RNA copies does not statistically decrease until day 8 from symptom onset. After day 8, exhaled viral RNA levels dropped near the threshold of detection, and in many cases to undetectable levels. Very low levels of viral RNA were intermittently detected up to 20 days from symptom onset, with occasional spikes in a few participants (**Fig 2A**). These findings suggest that in outpatients with a low rate of co-morbidities (**Table 1**), viral loads on breath are substantially decreased after day 8 from the onset of symptoms.

**Fig 2.**
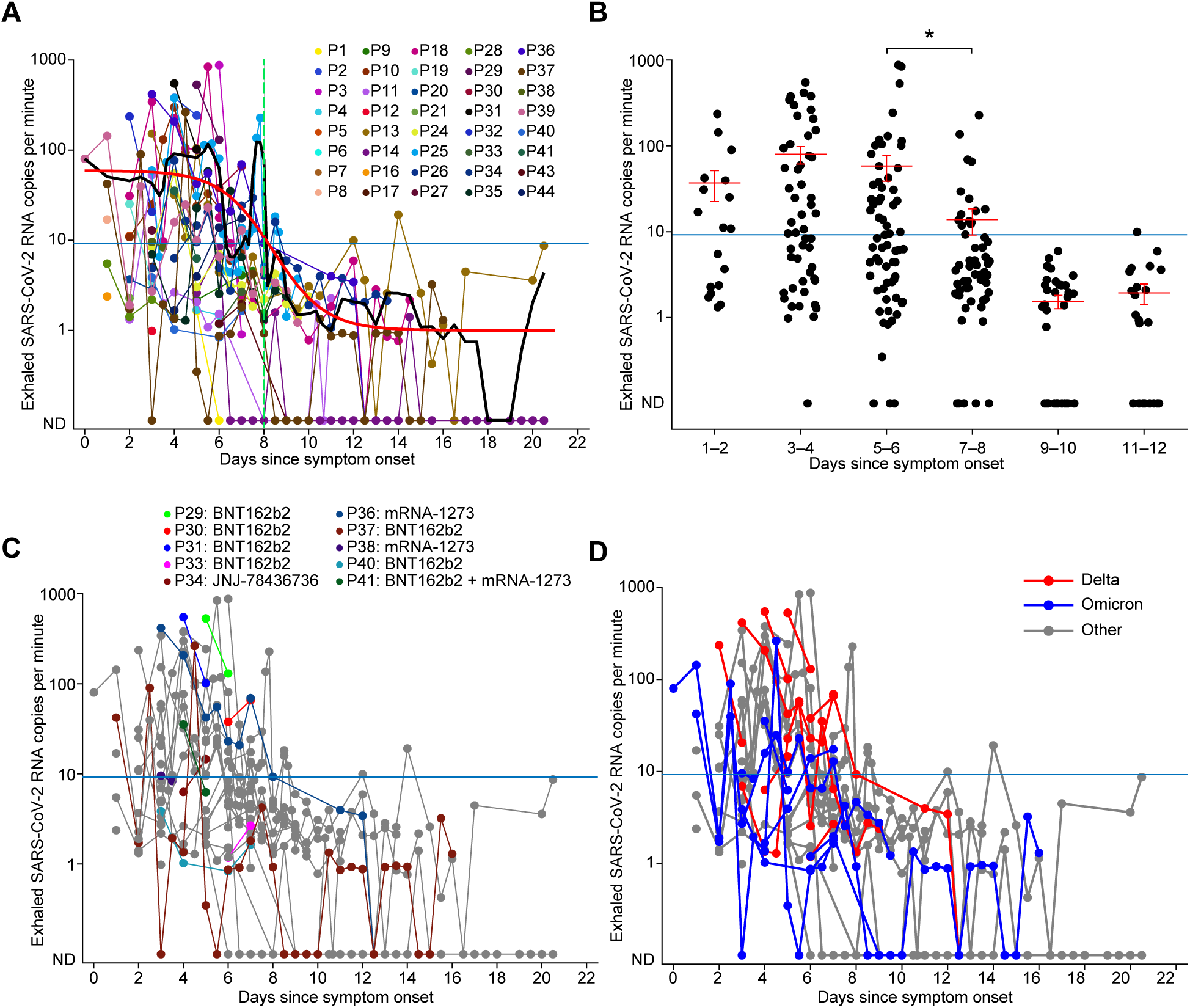
(A) Levels of exhaled SARS-CoV-2 RNA copies per minute over the course of infection. Each dot represents the daily average number of exhaled SARS-CoV-2 RNA copies exhaled per minute, for each participant. The overlaid black line represents the rolling average of exhaled viral RNA copies over days since symptom onset. The overlaid red line is the fitted S curve (R^2^ = 0.7657, adjusted R^2^ = 0.7567, RMSE: 0.3755). The green vertical dashed line indicates the inflection point of the S curve. (B) Viral RNA copy numbers binned over days since symptom onset. Each dot represents results from one sample. Mean and standard error are represented by red lines within each bin. * indicates *P* < 0.05. (C) The same data from panel A are plotted, with colored dots representing samples that were collected from vaccinated participants, and grey dots representing unvaccinated participants. (D) The same data from panel A are plotted with blue indicating participants infected with Omicron strains; red indicating infection with Delta strains; and grey indicating infections prior to Delta. All panels: blue horizontal line represents 35 C_T_ equivalent.

#### Vaccinated and unvaccinated individuals exhaled similar levels of SARS-CoV-2 RNA

Whether vaccinated individuals who experience breakthrough infection with COVID-19 shed lower levels of virus on breath is unknown. Our data set included 57 samples collected by 11 vaccinated participants with breakthrough infections (**Table 1**). We found that vaccinated and unvaccinated participants exhaled similar numbers of SARS-CoV-2 RNA copies (**Fig 2C**), when accounting for age, sex, presence of co-morbidities, days since symptom onset and symptom severity (Supplementary Appendix, Wilcoxon rank sum test, *P* = 0.31, z = 1.02) (**Fig 3A**). Viral RNA levels in 57 samples collected by vaccinated individuals on days 1 to 16 from symptom onset ranged from 0 to 549 exhaled copies per minute (%95 CI: [17, 78]) while viral loads for unvaccinated individuals on days 1 to 16 ranged from 0 to 876 exhaled copies per minute (%95 CI: [25, 55]).

**Fig 3.**
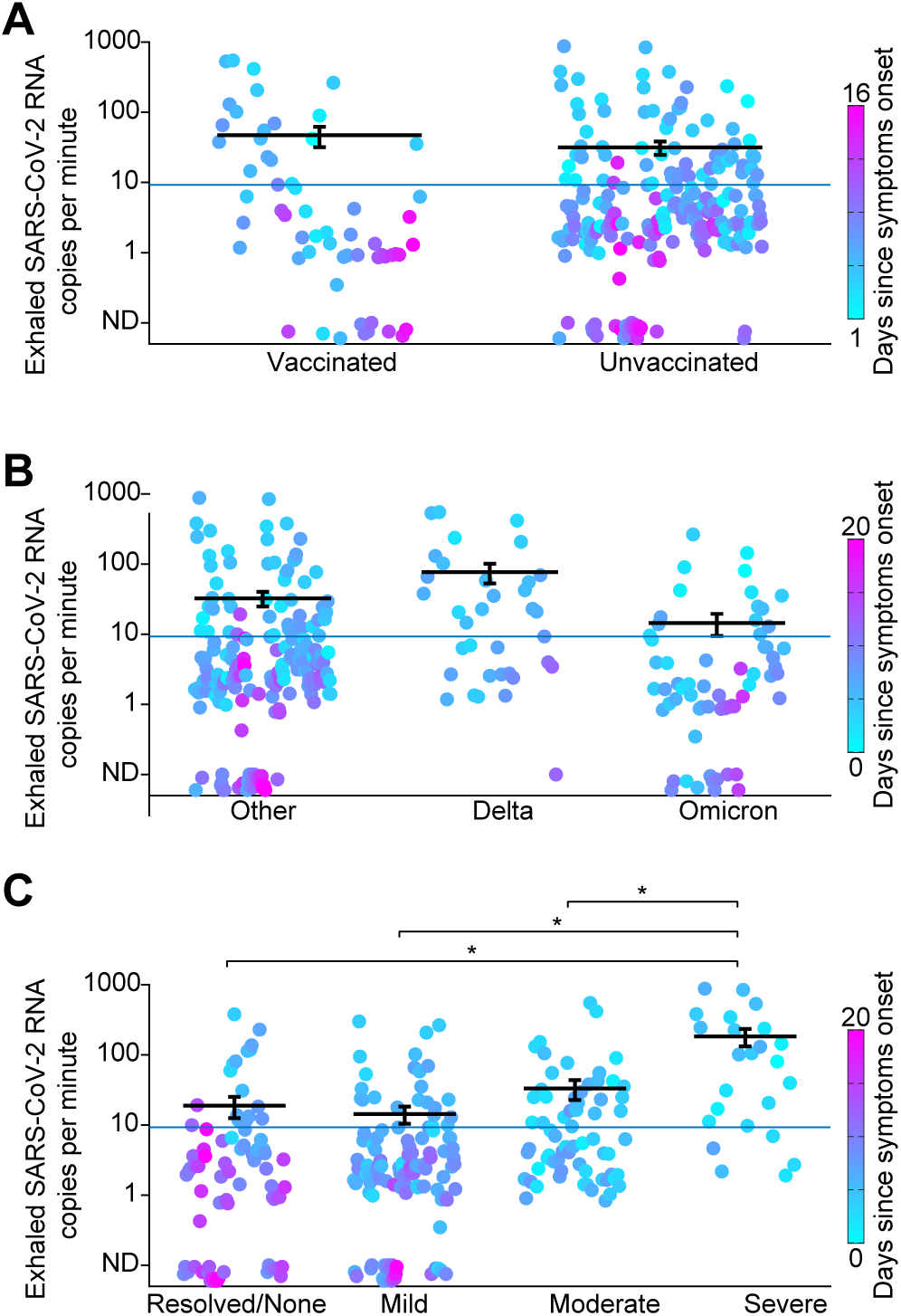
Levels of SARS-CoV-2 RNA copies in exhaled breath for (A) vaccination status, (B) viral variants and (C) self-reported symptom severity. Each colored dot represents one sample, and the color scale represents days since symptom onset. Black bars denote average, error bars denote standard error, and blue horizontal line represents 35 C_T_ equivalent. * indicates *P* < 0.05.

#### Levels of exhaled SARS-CoV-2 RNA were similar across viral variants

Whether infection by different viral variants impacts levels of virus shed on breath is poorly understood. In particular, whether infection with later SARS-CoV-2 variants considered to be more transmissible, such as Delta and Omicron, produces higher levels of viral shedding on breath than earlier variants is unknown. Genome sequencing was performed on viral RNA extracted from 9 exhaled breath condensate samples from 7 participants in order to determine SARS-CoV-2 viral clade (**Fig 1C**). For all other participants, clade was putatively assigned based on the variant prevalence in Chicago over a rolling 7-day average at time of symptom onset (n = 35) or date of diagnosis (n = 2; P15 and P42) (**Fig 1C**). Our data set included 194 samples from individuals infected by early SARS-CoV-2 variants between April 2020 and March 2021 (clades 20A, 20B, 20C, and 20G), 35 samples from individuals infected by Delta between July 2021 and October 2021 (clade 21J), and 65 samples from individuals infected by Omicron in December 2021 and January 2022 (clade 21K) (**Fig 1C**, with additional detail in the online data supplement **Table S1**). Levels of viral shedding on breath did not differ between these three groups when accounting for age, sex, presence of co-morbidities, and days since symptom onset (Tukey’s test, *P* > 0.05) (**Fig 3B**). Viral loads on breath in individuals infected with the early variants ranged from 0 to 876 RNA copies exhaled per minute (%95 CI: [17, 48]), while viral loads in individuals infected with the Delta variant ranged from 0 to 549 RNA copies exhaled per minute (%95 CI: [28, 126]), and viral loads for individuals infected with the Omicron variant ranged from 0 to 264 RNA copies exhaled per minute (%95 CI: [4, 25]) (**Fig 2D**). In a second confirmatory analysis, we compared peak levels of viral shedding on breath over infection course in individuals infected with each variant. Peak levels of viral shedding on breath were similar across variants (One-way ANOVA; F_2,41_ = 2.56, *P* = 0.089) (**Fig 1D**).

#### Exhaled SARS-CoV-2 RNA levels increased with self-reported symptom severity

Relationships between viral load and symptom severity have been studied using internal swabs to estimate levels of viral RNA (9, 38). However, whether exhaled SARS-CoV-2 viral loads correlate with symptom severity has been unexplored. We examined the number of SARS-CoV-2 RNA copies exhaled per minute across self-reported symptom severity groups (group: (mean [%95 CI]); Resolved/None: 19 [6, 31]; Mild: 14 [6, 22]; Moderate 33 [12, 53]; Severe: 183 [76, 289]) (**Fig 3C**), accounting for age, sex, presence of co-morbidities, and days since symptom onset. On average, exhaled viral RNA copies were significantly higher in samples collected when participants reported that their symptoms were Severe compared to those collected when symptoms were reported as Resolved/None, Mild, or Moderate (Tukey test; Resolved/None: *P* = 0.002, Mild: *P* < 0.001, or Moderate: *P* = 0.005). However, there was considerable individual variation reflected by large confidence intervals, and as can be seen in **Fig 3C**. For example, some individuals who were asymptomatic at the time of specimen collection were exhaling around 400 viral copies per minute (**Fig 3C**, left-most column), highlighting the importance of understanding the factors that contribute to individual variation in levels of viral shedding on breath (39).

## DISCUSSION

Our study used a newly-developed portable exhaled breath condensate collection device to capture breath specimens from COVID-19-tested outpatient participants. We used this data to characterize the dynamics of viral load on exhaled breath from the onset of symptoms through day 20. We found that COVID-19 infected participants exhaled an average of 80 viral RNA copies per minute during the first 8 days of infection, with significant variability, including peaks as high as 876 copies per minute. The average amount of viral RNA being exhaled into the environment did not statistically decrease until day 8 following symptom onset, when we found a steep drop to levels near the limit of detection, which persisted for up to 20 days from symptom onset, and included occasional spikes in a few participants. We further found that participants who rated their symptoms as severe were exhaling significantly more viral RNA copies per minute than participants who rated their symptoms as mild or moderate, though with high individual variance. We found no impact of vaccination status, viral variant, age, sex, or presence of co-morbidities.

Given the long-term effects of COVID-19 infection (30–32), and the potential cumulative risks of re-infection (33), reducing the burden on society due to COVID-19 will require strategies for infection and re-infection prevention (34); characterization of the dynamics of exhaled viral loads—especially during natural, unlabored breathing—is important for better understanding of the risk of onward infection.

Our study has several limitations. First, we had a small number (n = 11) of vaccinated participants, most of whom received the same vaccine (BNT162b2, n = 7), limiting our ability to measure the effect of vaccination on viral shedding on breath. Notably, because vaccines became available later in our study period, our vaccinated participants were likely infected with a later-emerging variant of SARS-CoV-2, potentially impacting comparisons with earlier participants (17). That said, we did not find any differences in exhaled viral loads across viral variants in this study. Second, self-reported symptom severity data is variable and adds uncertainty to those findings. While a majority of participants (95%) were in an outpatient setting, assessing severity through the use of defined clinical severity scores would have yielded a more reliable measure.

Despite these limitations, our findings make a significant contribution to understanding the dynamics of COVID-19 infection in an outpatient population. The majority of research on the dynamics viral load have focused on hospitalized inpatient populations, typically characterizing viral dynamics from day of diagnosis, relative to clinically-assessed disease severity. In contrast, relatively little is known about viral dynamics among outpatients from time of symptom onset, relative to symptom severity (9), and dynamics of viral load on breath over the course of infection are poorly understood. Most COVID-19 infections are experienced as outpatients in household and community settings. Therefore, in order to inform community transmission mitigation strategies—the environment in which the vast majority of transmission occurs—we need to understand the dynamics of the virus in an outpatient population. Significant effort has gone into characterizing viral shedding dynamics, which has led to the development and improvement of models of transmission and methods of infection mitigation. Notably, however, the lack of congruency in viral load dynamics between different swab locations within individuals erodes confidence in respiratory tract viral loads as indicators of infectiousness (39). Here we add a new element to this effort by directly characterizing viral shedding on breath, the major vector of transmission of COVID-19.

Our findings can be used to demonstrate future feasibility of estimating the amount of time it takes for an individual to exhale an infectious dose of SARS-CoV-2: For example, assuming a conservative estimate of infectious dose for COVID-19 of 300 virions (35, 36), a person exhaling 900 viral RNA copies per minute (as did peak shedders in our study) could be conservatively estimated to exhale an infectious dose in around 20 seconds (not safe for an elevator ride), whereas a person who is exhaling 10 viral copies per minute could be estimated to exhale an infectious dose in over 30 minutes (probably ok for an elevator ride) (37). Along these same lines, once the relationship between exhaled viral RNA and replication-competent virus is established, if combined with rapid SARS-CoV-2 detection methods, our device could be used to assist in development of an at-home infectiousness test.

Understanding the dynamics of exhaled viral loads during natural breathing over the course of infection is an important step toward understanding onward of SARS-CoV-2. Our findings support the possibility that an individual becomes less likely to infect others through breath beyond day 8 after symptom onset, though further work is needed to confirm that infectivity parallels viral RNA levels on breath. Notably, it is unknown why some patients exhale large amounts of viral loads and others do not. The method used here exploits a low-cost, portable, disposable, and straightforward-to-use device, allowing participants to complete multiple, unsupervised, at-home sample collection sessions after a single brief training session. This method has the potential to pave the way for large-scale, individualized research into viral shedding on breath for COVID-19, and for other existing and yet-to-come respiratory pathogens.

## MATERIALS AND METHODS

### Cohort Description

This was a prospective observational study of participants recruited from the Northwestern Memorial Hospital (NMH) Emergency Department (ED) and Immediate Care Clinics (ICCs) in Chicago, Illinois from April 27, 2020 until January 8, 2022. Participants were identified after an order for COVID-19 testing was placed by the clinical care team and they were recruited by study staff as a convenience sample. Any adult (at least 18 years) undergoing COVID-19 testing in the NMH ED or ICCs was eligible for study enrollment regardless of symptom severity. In general, the COVID-19 testing procedure in the clinic included collection of a nasopharyngeal swab and analysis of the direct swab using Abbott ID NOW, a qualitative rapid non-PCR nucleic acid-based test which provides binary results (positive or negative). COVID-19-positive participants had a median age of 34 years (range 20 to 62 years) and 45.45% were female (**Table 1**). All participants were outpatients, with the exception of 2 ED patients who were admitted. All participants provided voluntary informed consent, and all research procedures were approved by the Northwestern University IRB, Study #STU00212292.

### Exhaled breath condensate collection device

EBCD is a simple device that can be built for less than US $5.00 using readily available parts (**Fig S1**). It consists of a modified 50 CC catheter-tip syringe body and plunger, a plastic mouth piece (or nose piece), a one-way flutter valve/saliva trap and a cooling sleeve with insulating cover (**Fig S1A**). To assemble the device, the one-way valve is placed inside the mouth piece, oriented to prevent the participant from inhaling room air through the tube. The mouthpiece assembly is then placed into the wide end of the syringe body. The cooling sleeve with insulating cover is placed over the syringe body. To produce a sample, the participant simply breathes naturally and exhales through the EBCD for 10 minutes. As the participant exhales through the device, any saliva that may enter the tube is trapped by the valve casing while moisture in warm exhaled breath condenses on the cooled inner wall of the device, producing a liquid condensate sample (**Fig S1B**). After the breathing session, the valve and mouth piece are removed, the plunger is inserted, and the liquid sample is plunged into a vial, which contains 1 mL of transport medium (**Fig S1C**). The resulting sample can be mailed to the lab for analysis. A typical 10-minute breathing session yields around 1 mL of breath condensate.

The EBCD has oral and nasal breath collection configurations (**Fig S2**). For this study, because mouth pieces are cheaper and more readily commercially available, the device was configured with a mouth piece and participants provided oral breathing samples. Because we were interested in SARS-CoV-2 RNA levels in exhaled breath during natural breathing as a baseline condition of potential viral transmission, we instructed participants to breathe at a natural pace, and avoid talking, shouting, singing, or any other exertive breathing. Natural breathing should produce condensate samples with minimal liquid saliva contamination: Liquid saliva is distinct from saliva which is volatilized naturally during exhale; volatilized saliva is an expected constituent of our sample, while liquid saliva is a contaminant which can enter the sample via spitting or drooling. To avoid saliva contamination via drooling or spittle, the configuration of the mouthpiece and one-way valve were designed to prevent spitting or drooling into the device and to trap any minimal saliva that enters the mouthpiece as a liquid. We confirmed the efficacy of this design in eliminating liquid saliva contamination by performing viscosity measurements on control samples using an rheometer (Anton Paar MCR302) equipped with a Peltier temperature controller set to 25°C and a CP50-1 cone and plate fixture. If oral samples collected with our device are contaminated by saliva, we would expect oral samples to have higher viscosity compared to nasal samples. Two healthy participants produced 3 samples each: nasal and oral breath samples using the EBCD, and saliva samples by spitting into a sample tube, resulting in 6 samples. 0.6 mL of each sample was added to the Peltier stage at 25°C, and was subjected to a shear rate sweep from 5 to 500 s^-1^ at 5 s^-1^ increment. Three measurements were performed on each sample. MilliQ water was also measured for comparison. We found that nasal breath, oral breath and water were indistinguishable in terms of viscosity, with saliva having much higher viscosity (**Fig S2**). Thus EBCD oral breath samples were not contaminated by saliva.

### Specimen collection

Participants were asked to provide an initial breath sample either in the clinic, from home following in-person study-team instruction on how to use the EBCD device, or from home following study-team instruction provided remotely over videoconferencing software or via online video. Participants were then invited to participate in voluntary longitudinal home collection of additional exhaled breath samples, typically two per day, for variable duration ranging from 2 to 20 days. Participant demographics were collected at baseline along with the date of symptom onset as determined by clinical notes and participant interviews. At the time of each sample collection, participants recorded time of day, symptoms, and subjective self-assessed overall symptom severity (resolved/none, mild, moderate, severe). Exhaled breath condensate samples were taken from 60 participants within an average of 19 hours (range 1–120, %95 CI: [13.3, 25.1]) after clinical diagnostic COVID-19 testing. A subset of 31 COVID-19-positive participants (70.5%) produced an additional 250 samples longitudinally from home (range 2–30, %95 CI: [6, 12]). In total, 60 participants produced 312 samples, two of which were unusable due to accidental spillage, leaving 310 samples for analysis.

### Sample analysis

Samples were collected into vials containing 1 mL molecular transport medium (Primestore, Longhorn Vaccines and Diagnostics) and stored at 4°C. For analysis, the full volume of each sample, representing 10 minutes of natural breathing, was concentrated to a final volume of 200 µL using Amicon Ultra filters (3 kDa). RNA was extracted using the QIAamp MinElute virus spin kit using the full concentrated volume of each sample as an input, and using a 40 µL elution. Reverse-Transcriptase quantitative Polymerase Chain Reaction (RT-qPCR) was performed in triplicate, using TaqMan probes and N1-targeted primers (IDT 2019-nCoV kit, catalog #10006606) following the CDC 2019-Novel Coronavirus (2019-nCoV) Real-Time RT-PCR Diagnostic Panel (CDC-006-00019). Cycle threshold (C_T_) values were obtained using ThermoFisher QuantStudio Real-Time PCR software. The estimated number of viral copies represented by the C_T_ was determined using a standard curve generated with a dilution series of the 2019-nCoV_N_positive control plasmid (IDT catalog #10006625) (**Fig 1B**). To convert C_T_ value to the number of viral copies, we used Matlab’s polyfit function to fit a first-degree polynomial to the standard data. The data were modeled as *y* = *a* ∗ log(*x*) + *b*, where *y* is the C_T_ value, *x* is the number of viral copies, *a* and *b* and are the polynomial coefficients (*a* = – 3.63, *b* = 38.51). Use of the full volume of sample collected over a 10-minute breathing session normalized all samples to represent 10 minutes of natural breathing. This allowed us to convert C_T_ values to numbers of viral copies that were exhaled per minute over a ten-minute breathing session.

### Sequencing library preparation, Illumina sequencing, and genome assembly

Complementary DNA (cDNA) synthesis was performed with the SuperScript IV First Strand Synthesis Kit (Thermo) using random hexamer primers according to manufacturer’s specifications. Direct amplification of viral genome DNA was performed as previously described using the Artic Network version 4 primers. Sequencing library preparation of genome amplicon pools was performed using the SeqWell plexWell 384 kit per manufacturer’s instructions. Pooled libraries of up to 96 genomes were sequenced on the Illumina MiSeq using the V2 500 cycle kit. Sequencing reads were trimmed to remove adapters and low-quality sequences using Trimmomatic v0.36. Trimmed reads were aligned to the reference genome sequence of SARS-CoV-2 (accession MN908947.3) using bwa v0.7.15. Pileups were generated from the alignment using samtools v1.9 and consensus sequence determined using iVar v1.2.2 with a minimum depth of 10, a minimum base quality score of 20, and a consensus frequency threshold of 0 (i.e., majority base as the consensus). SARS-CoV-2 clades were assessed using Nextclade (clades.nextstrain.org) and Pango lineages were assigned to the consensus sequences using pangolin software.

### Putative clade designations

After determining viral clade for 7 participants by sequencing, we assigned a putative clade to each remaining participant based on time-matched SARS-CoV-2 sequencing data. We downloaded all available SARS-CoV-2 genome data from Chicago between April 2020 and February 2021 (n = 8,155 genomes) from GISAID (https://www.gisaid.org/) as of July 27th, 2022 (Shu et al., 2017). Clade designations for sequences were determined using Nextclade’s phylogenetic nomenclature system (Aksamentov et al., 2021) and a 7-day rolling average was used to estimate clade frequency by date. Participants were assigned putative clade designations based on the most prevalent clade at the date of symptom onset (when available, n = 35) or date of diagnosis (n = 2; P15 and P42). We performed all calculations in base R and visualized the data using the R package ggplot2 ^33^.

### Statistical analysis

To examine the effect of self-reported symptom severity on exhaled viral loads, we used a generalized linear mixed model to control for age, sex, presence of co-morbidities, and days since symptom onset. To examine the effect of days since symptom onset on exhaled viral loads, we used a generalized linear mixed model to control for age, sex, presence of co-morbidities, and self-reported symptom severity. Subsequent analysis of variance was performed, and pair-wise comparisons between self-reported symptom groups (Resolved/None, Mild, Moderate, Severe) and between two-day bins of days since symptom onset (days 1–2, 3–4, 5–6, 7–8, 9–10, 11–12) were performed using Tukey’s Honest Significant Difference test on the estimated marginal means of each group or bin. Statistical analyses were performed on the logarithmic-transformed number of viral RNA copies. Not Detected (ND) values were set to zero and a constant (1) was added to all values prior to log transformation. Statistical significance threshold was *P* = 0.05. To determine the timing of reduction of viral shedding on breath, longitudinal data were modeled using Matlab’s *cftool* toolbox, which showed a best-fit to the S-curve model. The inflection point of the modeled curve was considered to represent the time of steepest drop in viral RNA copy numbers. We used a Wilcoxon rank sum test to examine the difference in exhaled viral RNA copies between vaccinated and unvaccinated participants. Details of statistical analysis are included in the Supplementary Methods.

## Data Availability

All data produced in the present study are available upon reasonable request to the authors

## ACKNOWLEDGEMENTS

Thanks to Goodwin A. Lane for extensive contributions in study planning and data collection, and thanks to Carol O’Dea and Samuel Newton. This research was also supported in part through the computational resources and staff contributions provided for the Quest high performance computing facility at Northwestern University, which is jointly supported by the Office of the Provost, the Office for Research, and Northwestern University Information Technology. Clinical data collection was supported in part by the Northwestern Medicine Enterprise Data Warehouse. This work was supported by the Northwestern University NUSeq Core Facility. Viscosity measurements were performed in the Analytical bioNanoTechnology Equipment Core Facility (ANTEC) of the Simpson Querrey Institute for BioNanotechnology at Northwestern University. ANTEC is currently supported by the Soft and Hybrid Nanotechnology Experimental (SHyNE) Resource (NSF ECCS-2025633) and Feinberg School of Medicine, Northwestern University. The Simpson Querrey Institute for BioNanotechnology, Northwestern University Office for Research, U.S. Army Research Office, and the U.S. Army Medical Research and Materiel Command have also provided funding to develop this facility. A special thanks to the participants whose willingness to participate during their illness made this work possible.

## AUTHOR CONTRIBUTIONS

Data Analysis: G.L., G.Z., J.F.H., L.S., R.L.-R., E.A.O., X.W., C.M.W., E.W., A.A., A.A., D.Y., R.A., C.Z. All authors contributed to the study design, and review and editing of the first draft manuscript.

## Supplementary Materials for

### This PDF file includes

Supplementary Text

Fig. S1

Fig. S2

Tables S1 to S8

References (1 to 2)

## Supplementary Text

### Cohort Description

This was a prospective observational study of participants recruited from the Northwestern Memorial Hospital (NMH) Emergency Department (ED) and Immediate Care Clinics (ICCs) in Chicago, Illinois from April 27, 2020 until January 8, 2022. Participants were identified after an order for COVID-19 testing was placed by the clinical care team and they were recruited by study staff as a convenience sample. Any adult (at least 18 years) undergoing COVID-19 testing in the NMH ED or ICCs was eligible for study enrollment regardless of symptom severity. In general, the COVID-19 testing procedure in the clinic included collection of a nasopharyngeal swab and analysis of the direct swab using Abbott ID NOW, a qualitative rapid non-PCR nucleic acid-based test which provides binary results (positive or negative). COVID-19-positive participants had a median age of 34 years (range 20 to 62 years) and 45.45% were female (**Table 1**). All participants were outpatients, with the exception of 2 ED patients who were admitted. All participants provided informed consent, and all research procedures were approved by the Northwestern University IRB, Study #STU00212292.

### Exhaled breath condensate collection device

Exhaled breath condensate samples were collected using the Exhaled Breath Collection Device (EBCD), developed by our lab. EBCD is a simple device that can be built for less than US $5.00 using readily available parts (**Fig S1**). It consists of a modified 50 CC catheter-tip syringe body and plunger, a plastic mouth piece, a one-way flutter valve/saliva trap and a cooling sleeve with insulating cover (**Fig S1A**). To assemble the device, the one-way valve/saliva trap is placed inside the mouth piece, oriented to trap saliva while preventing the participant from inhaling room air through the tube. The mouthpiece assembly is then placed into the wide end of the syringe body. The cooling sleeve with insulating cover is placed over the syringe body. To produce a sample, the participant simply breathes naturally and exhales through the EBCD for 10 minutes. As the participant exhales through the device, moisture in warm exhaled breath condenses on the cooled inner wall of the device, producing a liquid condensate sample (**Fig S1B**). After the breathing session, the valve and mouth piece are removed, the plunger is inserted, and the liquid sample is plunged into a vial, which contains 1 mL of transport medium (**Fig S1C**). The resulting sample can be mailed to the lab for analysis. A typical 10-minute breathing session yields around 1 mL of breath condensate.

### Specimen collection

Participants were asked to provide an initial breath sample either in the clinic, from home following in-person study-team instruction on how to use the EBCD device, or from home following study-team instruction provided remotely over videoconferencing software or via online video. Participants were then invited to participate in voluntary longitudinal home collection of additional exhaled breath samples, typically two per day, for variable duration ranging from 2 to 20 days. Participant demographics were collected at baseline along with the date of symptom onset as determined by clinical notes and participant interviews. At the time of each sample collection, participants recorded time of day, symptoms, and subjective self-assessed overall symptom severity (resolved/none, mild, moderate, severe). Exhaled breath condensate samples were taken from 60 participants within an average of 19 hours (range 1–120, %95 CI: [13, 25]) after clinical diagnostic COVID-19 testing (**Fig 1A**). A subset of 31 COVID-19-positive participants (70.5%) produced an additional 250 samples longitudinally from home (range 2–30, %95 CI: [6, 12]). In total, 60 participants produced 312 samples, two of which were unusable due to accidental spillage, leaving 310 samples for analysis.

### Detailed list of steps for assembly and use of Exhaled Breath Collection Device

1 Prepare device:
  - Trim syringe tip to a length of approximately 6 mm.
  - Remove plunger from syringe body.
  - Insert flutter valve inside the mouth piece until the mouthpiece hits the stop on the flutter valve, which should be oriented such that air will flow from the smaller end through the larger end of the mouthpiece.
  - Insert the assembled mouthpiece approximately 15 mm into the large end of the syringe body.
  - Roll a cooling sleeve (> 2 hrs at –20 °C) over the syringe tube starting from the tip end.
  - Cover the cooling sleeve with the insulating cover.
2 Produce a sample:
  - Seated comfortably, place the mouth piece into the mouth.
  - Breathing naturally over ten minutes, inhale through the nose and exhale through the mouth and thus through the device.
  - Over the course of the session, keep the device horizontal.
3 Recover a sample:
  - Keeping the device horizontal, remove the mouthpiece assembly and cooling sleeve, and insert the syringe plunger.
  - Orient the tip of the device into the collection vial, and using a slow and controlled motion, plunge the liquid sample into the collection vial.

### Sample analysis

Samples were collected into vials containing 1 mL molecular transport medium (Primestore, Longhorn Vaccines and Diagnostics) and stored at 4°C. For analysis, samples were concentrated to a final volume of 200 µL using Amicon Ultra filters (3 kDa). RNA was extracted using the QIAamp MinElute virus spin kit in a 40 µL elution. Reverse-Transcriptase quantitative Polymerase Chain Reaction (RT-qPCR) was performed in triplicate, using TaqMan probes and N1-targeted primers (IDT 2019-nCoV kit, catalog #10006606) following the CDC 2019-Novel Coronavirus (2019-nCoV) Real-Time RT-PCR Diagnostic Panel (CDC-006-00019). Cycle threshold (C_T_) values were obtained using ThermoFisher QuantStudio Real-Time PCR software. The estimated number of viral copies represented by the C_T_ was determined using a standard curve generated with a dilution series of the 2019-nCoV_N_positive control plasmid (IDT catalog #10006625) (**Fig 1B**). To convert C_T_ value to the number of viral copies, we used Matlab’s *polyfit* function to fit a first-degree polynomial to the standard data. The data were modeled as *y* = *a* ∗ log(*x*) + *b*, where *y* is the C_T_ value, *x* is the number of viral copies, and *a* and *b* are the polynomial coefficients (*a* = – 3.63, *b* = 38.51). This allowed us to convert C_T_ values to numbers of viral copies that were exhaled over a ten-minute breathing session.

### Putative clade designations

After determining viral clade for 7 participants by sequencing, we assigned a putative clade to each remaining participant based on time-matched SARS-CoV-2 sequencing data. We downloaded all available SARS-CoV-2 genome data from Chicago between April 2020 and February 2021 (n = 8,155 genomes) from GISAID (https://www.gisaid.org/) as of July 27th, 2022 (1). Clade designations for sequences were determined using Nextclade’s phylogenetic nomenclature system (1) and a 7-day rolling average was used to estimate clade frequency by date. Participants were assigned putative clade designations based on the most prevalent clade at the date of symptom onset (when available, n = 35) or date of diagnosis (n = 2; P15 and P42). We performed all calculations in base R and visualized the data using the R package ggplot2 (2).

### Statistics pertaining to Result 1: Average levels of SARS-CoV-2 RNA on breath decreased significantly around day 8 from symptom onset

To model the change of the number of viral copies across days since symptom onset, we fitted an S-Curve model to the average time course of viral RNA exhaled per minute. The average time course was smoothed using a moving average with a window of 3. The curve fitting was performed using Matlab’s *cftool* toolbox. The S-Curve model was *y* = *a*/(*b* + exp(*C* ∗ *x*)), where *y* is the log-transform of the number of viral RNA copies, *x* is the days since symptom onset, and *a*, *b*, and *c* are coefficients of the model. The inflection point of the modeled curve was considered to represent the time of steepest drop in viral RNA copies. The inflection point was calculated by first using the model parameters to find the fitted number of viral RNA copies from day 1 to day 20 with a step of 0.5 days, then calculating the point of maximal change from the fitted vector. The goodness of the model was qualified using R-squared and adjusted R-squared. We also compared the results to other models including Polynomial, Exponential, Fourier, Gaussian and Sum of Sine (**Table S2**).

### Statistics pertaining to Result 2: Exhaled SARS-CoV-2 RNA levels did not differ in 11 vaccinated individuals

The number of exhaled SARS-CoV-2 RNA copies for vaccinated participants ranged from 0 to 549 (Mean: 47, %95 CI: [17, 78]). A Wilcoxon rank-sum test suggested that the number of exhaled viral RNA copies for unvaccinated participants (Range 0**–**876, Mean: 32, %95 CI: [18, 45]) was not statistically significant different from that of vaccinated participants (Wilcoxon rank sum test; *P* = 0.30, z = 1.03). In a second confirmatory analysis, we used a generalized linear mixed model to control for potential confounds (age, sex, the presence of comorbidity, days since symptom onset and self-reported symptom severity), followed by a post-hoc Tukey test on the estimated marginal means (*P* = 0.75)

### Statistics pertaining to Result 3: Levels of exhaled SARS-CoV-2 RNA were similar across viral variants

To compare the number of exhaled SARS-CoV-2 RNA copies for samples infected by early SARS-CoV-2 variants between April 2020 and March 2021 (clades 20A, 20B, 20C, and 20G), Delta July 2021 and October 2021 (clade 21J), and Omicron in December 2021 and January 2022 (clade 21K), we used a generalized linear mixed model to control for potential confounds (age, sex, the presence of comorbidity, days since symptom onset and self-reported symptom severity). Post-hoc Tukey test revealed that the number of exhaled SARS-CoV-2 RNA copies were not statistically different across variants (*P* > 0.05; **Table S3, S4**)

### Statistics pertaining to Result 4: Exhaled SARS-CoV-2 RNA levels increased with self-reported symptom severity

To examine the effect of self-reported symptom severity on exhaled viral loads, we used a generalized linear mixed model to control for age, sex, the presence of co-morbidities, and days since symptom onset. Subsequent analysis of variance was performed, and pair-wise comparisons between symptom groups (Resolved/None, Mild, Moderate, Severe) were performed using Tukey’s Honest Significant Difference test on the estimated marginal means of each group. Statistical analyses were performed on the logarithmic-transformed number of viral RNA copies. Not Detected (ND) values were set to zero and a constant 1 added to all values prior to log transformation. Statistical significance threshold was *P* = 0.05. The ANOVA analysis revealed a statistically significant effect of self-reported symptom severity on the number of SARS-CoV-2 RNA copies exhaled per minute (F_3, 283_ = 14.76, *P* < 0.001). Post-hoc Tukey’s test using the estimated marginal mean suggested that the number of viral RNA copies exhaled were statistically higher when the participants reported that their symptoms were Severe, as compared to Resolved/None (*P* = 0.002), Mild (*P* < 0.001), or Moderate (*P* = 0.005) (**Table S5, S6**).

In a second confirmatory analysis to examine the effect of days since symptom onset on exhaled viral RNA loads, samples were binned according to their days from symptom onset (1–2, 3–4, 5– 6, 7–8, 9–10, 11–12 days). We used a generalized linear mixed model to control for age, sex, presence of co-morbidities, and self-reported symptom severity. Subsequent analysis of variance was performed, and pair-wise comparisons between bins was performed using Tukey’s Honest Significant Difference test on the estimated marginal means of each bin. Statistical analyses were performed on the logarithmic-transformed number of viral RNA copies. Not Detected (ND) values were set to zero and a constant 1 was added to all values prior to log transformation. Statistical significance threshold was *P* = 0.05. The mean viral RNA copies across bins were compared using ANOVA analysis, revealing a main effect of bins (F_5, 242_ = 11.15, *P* < 0.001). Levels were similar across bins including days 1 to 6, with significant decreases evident with the bin containing days 7 to 8 (Tukey’s test; *P* < 0.05; **Fig 2C**; **Tables S7, S8**).

---

**Figure S1:**
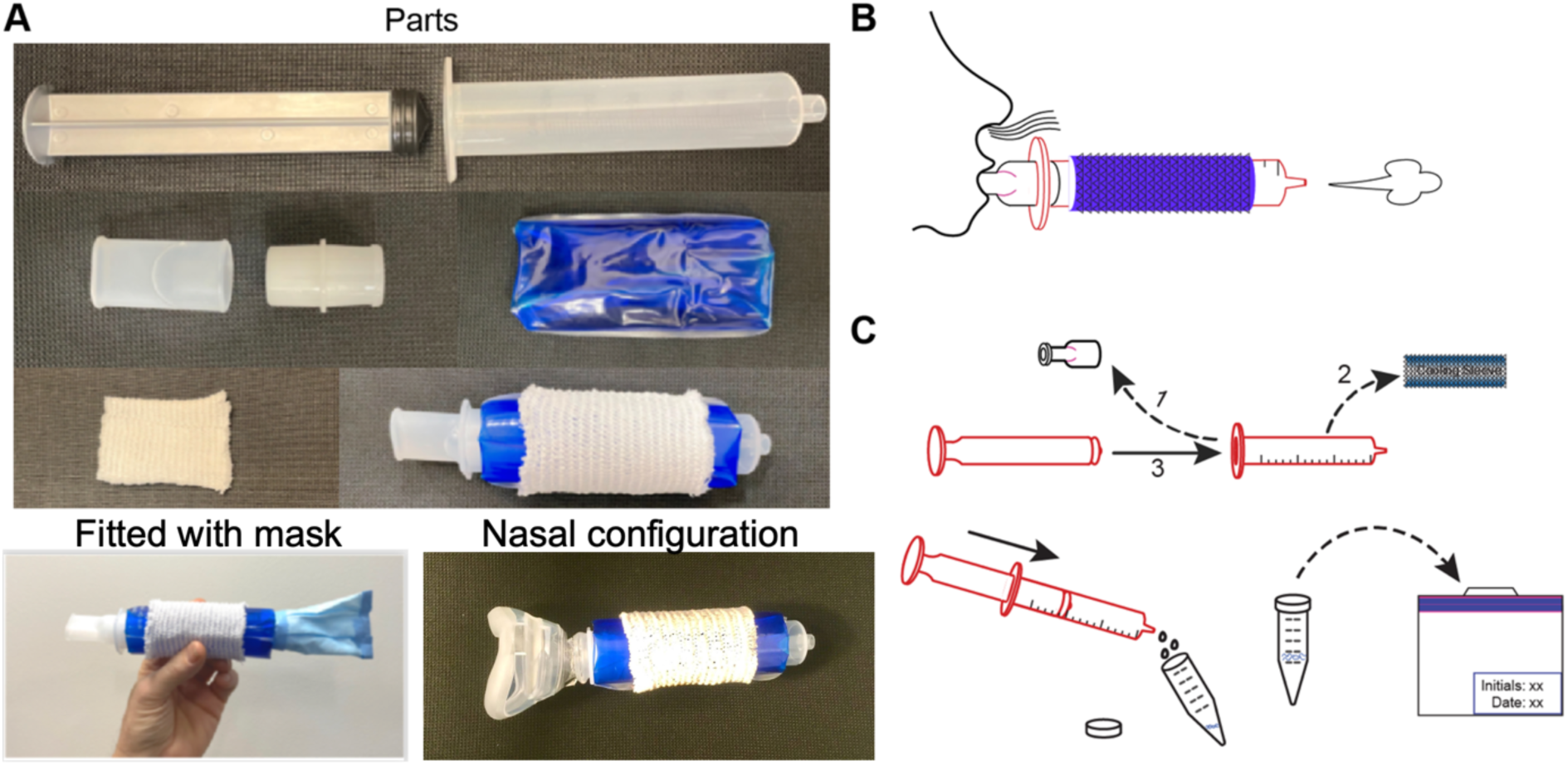
Exhaled Breath Collection Device Parts and Use. (A) Exhaled Breath Collection Device parts include a 50 CC catheter-tip syringe plunger and body, with tip trimmed to approximately 6 mm length (Monoject, Cardinal Health, top panel); a mouthpiece and one-way flutter valve (Vyaire, middle left); a cooling sleeve (Torex, middle right); and a cloth insulating cover (Surgilast, lower left). The assembled Exhaled Breath Collection Device is shown lower right; the device can be fitted with a mask cover over the narrow end by cutting a 10 cm square from a surgical mask and taping it over the end of the Exhaled Breath Collection Device (bottom) for use in clinical settings. (B) Exhaled Breath Collection Device in use. The user performs a breath condensate collection session by placing the mouthpiece in their mouth, breathing at a natural pace, and exhaling through the device at a natural resting pace. (C) Recovering a sample from the Exhaled Breath Collection Device. Once the breathing session is complete, the mouthpiece and cooling sleeve assemblies are removed, and the plunger is inserted in the wide end of the syringe body (top). The condensate sample is then recovered from the syringe body by pushing the plunger through the syringe body, forcing the liquid into a sample vial, which contains MTM (Primestore). Sample vials can be mailed for lab analysis (bottom).

**Figure S2.**
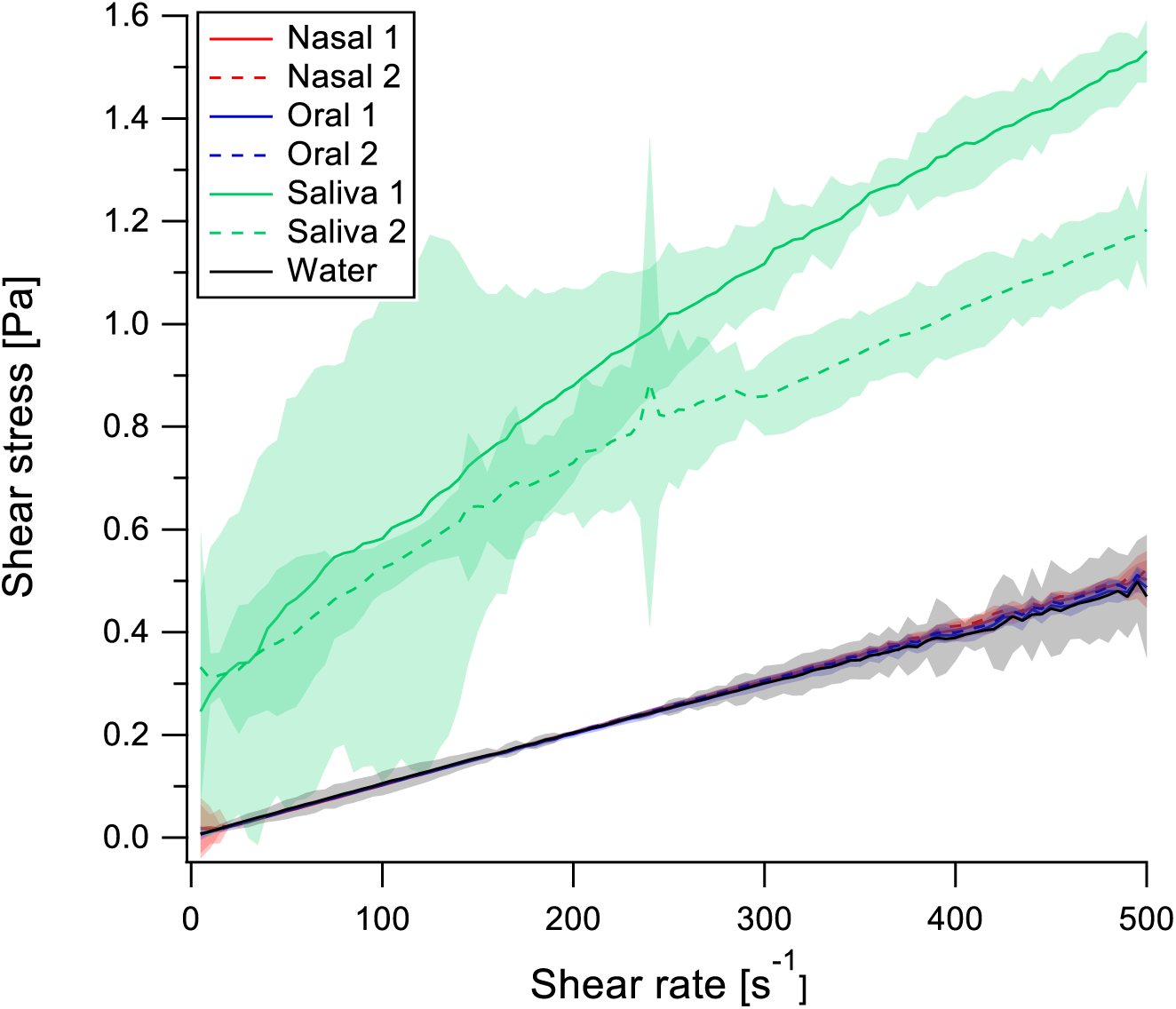
Shear rate vs. shear stress plots of condensates (nasal, red, oral, blue), saliva (green) and water (black), suggesting minimal contamination from saliva. Error shades correspond to 95% confidence levels. Nasal and oral samples were indistinguishable from each other and from water, strongly suggesting that oral breath samples collected with EBCD are not contaminated by saliva, and rather represent a pure exhaled breath condensate sample.

**Table S1.** Detail of viral clades, peak exhaled viral load, number of samples and days since symptom onset for each participant.

| Participant | Clade | Peak exhaled<br>SARS-CoV-2 RNA copies<br>per minute | Samples | Days since symptom<br>onset (min-max[mean]) |
| --- | --- | --- | --- | --- |
| P1 | 20C | 2 | 2 | 5–6[5.5] |
| P2 | 20C | 380 | 2 | 4–5[4.5] |
| P3 | 20C | 876 | 2 | 6–7[6.5] |
| P4 | 20C | 2 | 2 | 5–6[5.5] |
| P5 | 20C | 11 | 1 | 2–2[2.0] |
| P6 | 20C | 5 | 1 | 7–7[7.0] |
| P7 | 20A | Not Detected | 1 | 11–11[11.0] |
| P8 | 20C | 17 | 1 | 1–1[1.0] |
| P9 | 20C | 5 | 6 | 5–9[7.3] |
| P10 | 20A | 298 | 11 | 2–7[4.7] |
| P11 | 20A | 53 | 14 | 2–11[6.8] |
| P12 | 20A | 1 | 1 | 3–3[3.0] |
| P13 | 20B | 152 | 25 | 3–20[10.8] |
| P14 | 20A | 8 | 28 | 5–20[12.3] |
| P15 | 20C | 1 | 1 | * |
| P16 | 20G | 2 | 1 | 1–1[1.0] |
| P17 | 20C | 5 | 1 | 3–3[3.0] |
| P18 | 20G | 842 | 21 | 2–14[8.0] |
| P19 | 20G | 25 | 2 | 2–3[2.5] |
| P20 | 20G | 4 | 2 | 2–3[2.5] |
| P21 | 20G | 2 | 1 | 2–2[2.0] |
| P22 | 20G | 2 | 1 | * |
| P23 | 20G | 5 | 2 | * |
| P24 | 20G | 55 | 12 | 3–10[6.8] |
| P25 | 20G | 378 | 27 | 3–9[6.4] |
| P26 | 20G | 77 | 21 | 3–13[8.2] |
| P27 | 20G | 13 | 1 | 3–3[3.0] |
| P28 | 20G | 5 | 4 | 1–4[2.5] |
| P29 | 21J | 532 | 2 | 5–6[5.5] |
| P30 | 21J | 66 | 2 | 6–7[6.5] |
| P31 | 21J | 549 | 2 | 4–5[4.5] |
| P32 | 21J | 236 | 2 | 2–3[2.5] |
| P33 | 21J | 3 | 2 | 6–7[6.5] |
| P34 | 21J | 15 | 2 | 4–5[4.5] |
| P35 | 21J | 58 | 12 | 3–9[6.0] |
| P36 | 21J | 416 | 11 | 3–12[7.2] |
| P37 | 21K | 264 | 30 | 1–16[8.5] |
| P38 | 21K | 10 | 2 | 3–3[3.0] |
| P39 | 21K | 144 | 17 | 0–9[5.1] |
| P40 | 21K | 4 | 4 | 3–7[5.0] |
| P41 | 21K | 35 | 2 | 4–5[4.5] |
| P42 | 21K | 1 | 2 | * |
| P43 | 21K | 2 | 4 | 6–10[8.0] |
| P44 | 21K | 17 | 4 | 4–7[5.5] |

\*Unavailable

**Table S2.**
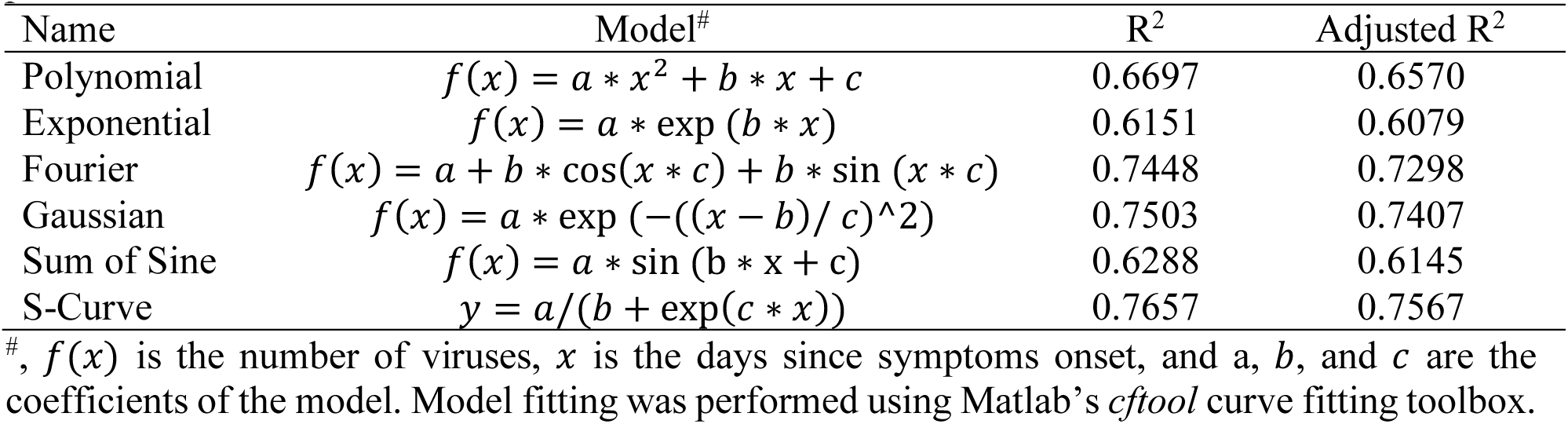
Comparison of different models fitted to the average time course of viral RNA exhaled per minute.

**Table S3.**
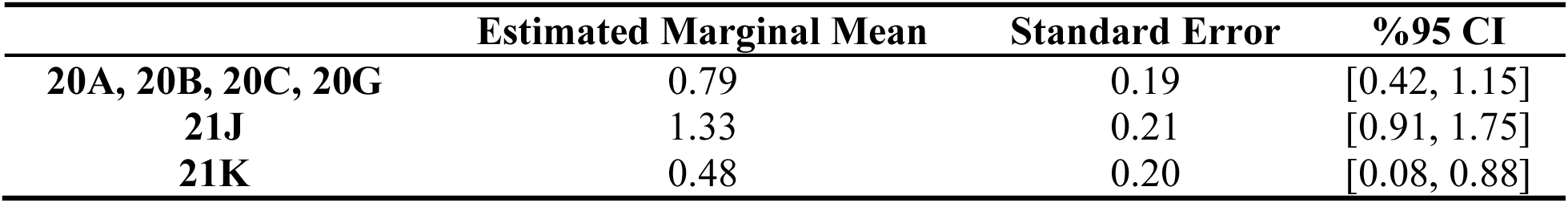
Estimated marginal mean of the number of SARS-CoV-2 RNA copies (log-transformed) for each of the variant.

|  | **Estimated Marginal Mean** | **Standard Error** | **%95 CI** |
| --- | --- | --- | --- |
| **20A, 20B, 20C, 20G** | 0.79 | 0.19 | [0.42, 1.15] |
| **21J** | 1.33 | 0.21 | [0.91, 1.75] |
| **21K** | 0.48 | 0.20 | [0.08, 0.88] |

**Table S4.**
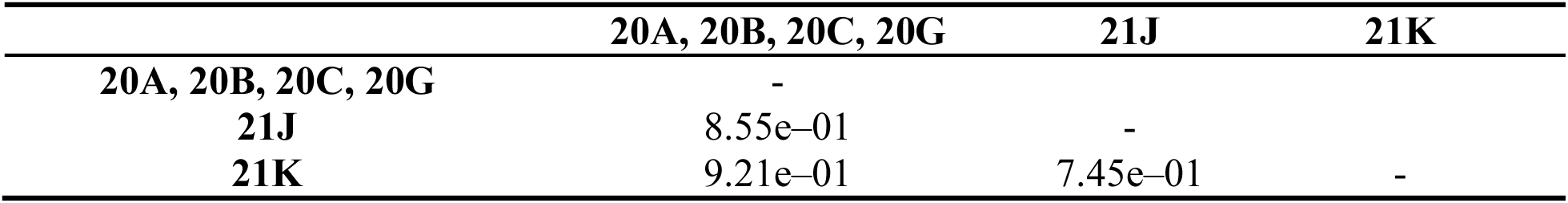
*P* values of post-hoc Tukey test on the number of SARS-CoV-2 RNA copies across variants.

**Table S5.** Estimated marginal mean of the number of SARS-CoV-2 RNA copies (log-transformed) for each of the symptom severity groups.

| **Symptom Severity** | **Estimated Marginal Mean** | **Standard Error** | **%95 CI** |
| --- | --- | --- | --- |
| **Resolved/None** | 0.87 | 0.11 | [0.66, 1.09] |
| **Mild** | 0.60 | 0.08 | [0.44, 0.76] |
| **Moderate** | 0.78 | 0.10 | [0.58, 0.98] |
| **Severe** | 1.39 | 0.13 | [1.13, 1.65] |

**Table S6.** *P* values of post-hoc Tukey test on the number of SARS-CoV-2 RNA copies across self-reported symptom severity.

| **Symptom Severity** | **Resolved/None** | **Mild** | **Moderate** | **Severe** |
| --- | --- | --- | --- | --- |
| **Resolved/None** | - |  |  |  |
| **Mild** | 7.80e-02 | - |  |  |
| **Moderate** | 8.85e-01 | 4.42e-01 | - |  |
| **Severe** | 2.21e-02* | 3.13e-05* | 4.93e-03* | - |

\* $P < 0.05$ .

**Table S7.** Estimated marginal mean of the number of SARS-CoV-2 RNA copies (log-transformed) for each of bin of days.

|  | **Estimated Marginal Mean** | **Standard Error** | **%95 CI** |
| --- | --- | --- | --- |
| **Day 1–2** | 1.12 | 0.14 | [0.84, 1.40] |
| **Day 3–4** | 1.35 | 0.09 | [1.17, 1.54] |
| **Day 5–6** | 1.19 | 0.09 | [1.01, 1.37] |
| **Day 7–8** | 0.90 | 0.10 | [0.70, 1.10] |
| **Day 9–10** | 0.53 | 0.13 | [0.28, 0.78] |
| **Day 11–12** | 0.45 | 0.15 | [0.16, 0.75] |

**Table S8.** *P* values of post-hoc Tukey test on the number of SARS-CoV-2 RNA copies across days since symptom onset.

|  | **Day 1–2** | **Day 3–4** | **Day 5–6** | **Day 7–8** | **Day 9–10** | **Day 11–12** |
| --- | --- | --- | --- | --- | --- | --- |
| **Day 1–2** | - |  |  |  |  |  |
| **Day 3–4** | 5.87e–01 | - |  |  |  |  |
| **Day 5–6** | 9.95e–01 | 5.72e–01 | - |  |  |  |
| **Day 7–8** | 6.54e–01 | 1.20e–04* | 2.71e–02* | - |  |  |
| **Day 9–10** | 2.45e–03* | 4.38e–11* | 7.23e–08* | 1.94e–02* | - |  |
| **Day 11–12** | 1.32e–03* | 4.95e–10* | 2.86e–07* | 1.15e–02* | 9.94e–01 | - |

\* $P < 0.05$ .

## REFERENCES

1. Greenhalgh T, Jimenez JL, Prather KA, Tufekci Z, Fisman D, Schooley R. Ten scientific reasons in support of airborne transmission of SARS-CoV-2. Lancet 2021;397:1603–1605.

2. Bourouiba L. Turbulent gas clouds and respiratory pathogen emissions: Potential implications for reducing transmission of COVID-19. JAMA 2020;323:1837–1838.

3. Coleman KK, Tay DJW, Tan K Sen, Ong SWX, Than TS, Koh MH, Chin YQ, Nasir H, Mak TM, Chu JJH, Milton DK, Chow VTK, Tambyah PA, Chen M, Tham KW. Viral load of severe acute respiratory syndrome coronavirus 2 (SARS-CoV-2) in respiratory aerosols emitted by patients with coronavirus disease 2019 (COVID-19) while breathing, talking, and singing. Clin Infect Dis 2022;74:1722–1728.

4. Liu Y, Ning Z, Chen Y, Guo M, Liu Y, Gali NK, Sun L, Duan Y, Cai J, Westerdahl D, Liu X, Xu K, Ho K, Kan H, Fu Q, Lan K. Aerodynamic analysis of SARS-CoV-2 in two Wuhan hospitals. Nature 2020;582:557–560.

5. Scheuch G. Breathing is enough: For the spread of influenza virus and SARS-CoV-2 by breathing only. J Aerosol Med Pulm Drug Deliv 2020;33:230–234.

6. Stadnytskyi V, Bax CE, Bax A, Anfinrud P. The airborne lifetime of small speech droplets and their potential importance in SARS-CoV-2 transmission. Proc Natl Acad Sci 2020;117:11875–11877.

7. van Doremalen N, Bushmaker T, Morris DH, Holbrook MG, Gamble A, Williamson BN, Tamin A, Harcourt JL, Thornburg NJ, Gerber SI, Lloyd-Smith JO, de Wit E, Munster VJ. Aerosol and surface stability of SARS-CoV-2 as compared with SARS-CoV-1. N Engl J Med 2020;382:1564–1567.

8. He X, Lau EHY, Wu P, Deng X, Wang J, Hao X, Lau YC, Wong JY, Guan Y, Tan X, Mo X, Chen Y, Liao B, Chen W, Hu F, Zhang Q, Zhong M, Wu Y, Zhao L, Zhang F, Cowling BJ, Li F, Leung GM. Temporal dynamics in viral shedding and transmissibility of COVID-19. Nat Med 2020;26:672–675.

9. Stankiewicz Karita HC, Dong TQ, Johnston C, Neuzil KM, Paasche-Orlow MK, Kissinger PJ, Bershteyn A, Thorpe LE, Deming M, Kottkamp A, Laufer M, Landovitz RJ, Luk A, Hoffman R, Roychoudhury P, Magaret CA, Greninger AL, Huang ML, Jerome KR, Wener M, Celum C, Chu HY, Baeten JM, Wald A, Barnabas R V., Brown ER. Trajectory of viral RNA load among persons with incident SARS-CoV-2 G614 infection (Wuhan Strain) in association with COVID-19 symptom onset and severity. JAMA Netw Open 2022;5:e2142796.

10. Jones TC, Biele G, Mühlemann B, Veith T, Schneider J, Beheim-Schwarzbach J, Bleicker T, Tesch J, Schmidt ML, Sander LE, Kurth F, Menzel P, Schwarzer R, Zuchowski M, Hofmann J, Krumbholz A, Stein A, Edelmann A, Corman VM, Drosten C. Estimating infectiousness throughout SARS-CoV-2 infection course. Science 2021;373:eabi5273.

11. Ke R, Martinez PP, Smith RL, Gibson LL, Achenbach CJ, McFall S, Qi C, Jacob J, Dembele E, Bundy C, Simons LM, Ozer EA, Hultquist JF, Lorenzo-Redondo R, Opdycke AK, Hawkins C, Murphy RL, Mirza A, Conte M, Gallagher N, Luo CH, Jarrett J, Conte A, Zhou R, Farjo M, Rendon G, Fields CJ, Wang L, Fredrickson R, et al. Longitudinal analysis of SARS-CoV-2 vaccine breakthrough infections reveals limited infectious virus shedding and restricted tissue distribution. Open Forum Infect Dis 2022;9:ofac192.

12. Fennelly KP. Particle sizes of infectious aerosols: implications for infection control. Lancet Respir Med 2020;8:914–924.

13. Klompas M, Baker MA, Rhee C. Airborne Transmission of SARS-CoV-2. JAMA 2020;324:441–442.

14. Malik M, Kunze A-C, Bahmer T, Herget-Rosenthal S, Kunze T. SARS-CoV-2: Viral loads of exhaled breath and oronasopharyngeal specimens in hospitalized patients with COVID-19. Int J Infect Dis 2021;110:105–110.

15. Riediker M, Morawska L. Low exhaled breath droplet formation may explain why children are poor SARS-CoV-2 transmitters. Aerosol Air Qual Res 2020;20:1513–1515.

16. Lednicky JA, Lauzardo M, Alam MM, Elbadry MA, Stephenson CJ, Gibson JC, Morris JG. Isolation of SARS-CoV-2 from the air in a car driven by a COVID patient with mild illness. Int J Infect Dis 2021;108:212–216.

17. Lai J, Coleman KK, Tai S-HS, German J, Hong F, Albert B, Esparza Y, Srikakulapu AK, Schanz M, Maldonado IS, Oertel M, Fadul N, Gold TL, Weston S, Mullins K, McPhaul KM, Frieman M, Milton DK. Evolution of SARS-CoV-2 shedding in exhaled breath aerosols. medRxiv [preprint] 2022 Aug 1 *Available from* 101101/2022072722278121

18. Feng B, Xu K, Gu S, Zheng S, Zou Q, Xu Y, Yu L, Lou F, Yu F, Jin T, Li Y, Sheng J, Yen H-L, Zhong Z, Wei J, Chen Y. Multi-route transmission potential of SARS-CoV-2 in healthcare facilities. J Hazard Mater 2021;402:123771.

19. Lednicky JA, Lauzardo M, Fan ZH, Jutla A, Tilly TB, Gangwar M, Usmani M, Shankar SN, Mohamed K, Eiguren-Fernandez A, Stephenson CJ, Alam MM, Elbadry MA, Loeb JC, Subramaniam K, Waltzek TB, Cherabuddi K, Morris JG, Wu C-Y. Viable SARS-CoV-2 in the air of a hospital room with COVID-19 patients. Int J Infect Dis 2020;100:476–482.

20. Leding C, Skov J, Uhrbrand K, Lisby JG, Hansen KP, Benfield T, Duncan LK. Detection of SARS-CoV-2 in exhaled breath from non-hospitalized COVID-19-infected individuals. Sci Rep 2022;12:11151.

21. Leung NHL, Chu DKW, Shiu EYC, Chan K-H, McDevitt JJ, Hau BJP, Yen H-L, Li Y, Ip DKM, Peiris JSM, Seto W-H, Leung GM, Milton DK, Cowling BJ. Respiratory virus shedding in exhaled breath and efficacy of face masks. Nat Med 2020;26:676–680.

22. Ma J, Qi X, Chen H, Li X, Zhang Z, Wang H, Sun L, Zhang L, Guo J, Morawska L, Grinshpun SA, Biswas P, Flagan RC, Yao M. Exhaled breath is a significant source of SARS-CoV-2 emission. medRxiv [preprint] 2022 Jun 2 *Available from* 101101/2020053120115154

23. Ryan DJ, Toomey S, Madden SF, Casey M, Breathnach OS, Morris PG, Grogan L, Branagan P, Costello RW, De Barra E, Hurley K, Gunaratnam C, McElvaney NG, OBrien ME, Sulaiman I, Morgan RK, Hennessy BT. Use of exhaled breath condensate (EBC) in the diagnosis of SARS-COV-2 (COVID-19). Thorax 2021;76:86–88.

24. Sawano M, Takeshita K, Ohno H, Oka H. RT-PCR diagnosis of COVID-19 from exhaled breath condensate: A clinical study. J Breath Res 2021;15:037103.

25. Verma R, Kim E, Degner N, Walter KS, Singh U, Andrews JR. Variation in severe acute respiratory syndrome coronavirus 2 bioaerosol production in exhaled breath. Open Forum Infect Dis 2022;9:ofab600.

26. Williams CM, Pan D, Decker J, Wisniewska A, Fletcher E, Sze S, Assadi S, Haigh R, Abdulwhhab M, Bird P, Holmes CW, Al-Taie A, Saleem B, Pan J, Garton NJ, Pareek M, Barer MR. Exhaled SARS-CoV-2 quantified by face-mask sampling in hospitalised patients with COVID-19. J Infect 2021;82:253–259.

27. Zhou L, Yao M, Zhang X, Hu B, Li X, Chen H, Zhang L, Liu Y, Du M, Sun B, Jiang Y, Zhou K, Hong J, Yu N, Ding Z, Xu Y, Hu M, Morawska L, Grinshpun SA, Biswas P, Flagan RC, Zhu B, Liu W, Zhang Y. Breath-, air-and surface-borne SARS-CoV-2 in hospitals. J Aerosol Sci 2021;152:105693.

28. Breslow NE, Clayton DG. Approximate inference in generalized linear mixed models. J Am Stat Assoc 2012;88:9–25.

29. Tukey JW. Comparing Individual Means in the Analysis of Variance. Biometrics 1949;5:99–114.

30. Kay LM. COVID-19 and olfactory dysfunction: a looming wave of dementia? J Neurophysiol 2022;128:436–444.

31. Graham EL, Clark JR, Orban ZS, Lim PH, Szymanski AL, Taylor C, DiBiase RM, Jia DT, Balabanov R, Ho SU, Batra A, Liotta EM, Koralnik IJ. Persistent neurologic symptoms and cognitive dysfunction in non-hospitalized Covid-19 “long haulers.” Ann Clin Transl Neurol 2021;8:1073–1085.

32. Xu E, Xie Y, Al-Aly Z. Long-term neurologic outcomes of COVID-19. Nat Med (In press).

33. AI-Aly Z, Bowe B, Xie Y. Outcomes of SARS-CoV-2 reinfection. Res Sq [preprint] 2022 Jun 17 *Available from* 1021203/rs3.rs-1749502/v1

34. Bohnert ASB, Bowling CB, Boyko EJ, Hynes DM, Ioannou GN, Iwashyna TJ, Maciejewski ML, O’hare AM, Viglianti EM. Burden of PCR-Confirmed SARS-CoV-2 reinfection in the U.S. veterans administration, March 2020 – January 2022. medRxiv *[preprint]* 2022 Mar 23 Availabel from 101101/2022032022272571

35. Karimzadeh S, Bhopal R, Nguyen Tien H. Review of infective dose, routes of transmission and outcome of COVID-19 caused by the SARS-COV-2: comparison with other respiratory viruses. Epidemiol Infect 2021;149:e96.

36. Prentiss M, Chu A, Berggren KK. Finding the infectious dose for COVID-19 by applying an airborne-transmission model to superspreader events. PLoS One 2022;17:e0265816.

37. van Rijn C, Somsen GA, Hofstra L, Dahhan G, Bem RA, Kooij S, Bonn D. Reducing aerosol transmission of SARS-CoV-2 in hospital elevators. Indoor Air 2020;30:1065–1066

38. Hakki S, Zhou J, Jonnerby J, Singanayagam A, Barnett J, Madon K, et al. Onset and window of SARS-CoV-2 infectiousness and temporal correlation with symptom onset: a prospective, longitudinal community cohort study. The Lancet Respiratory Medicine 2022; 10: 11: 1061–1073.

39. Ke R, Martinez P, Smith R, Gibson L, Mirza A, et al. Daily longitudinal sampling of SARS-CoV-2 infection reveals substantial heterogeneity in infectiousness. Nature Microbiology 2022; 7: 640–652.

## References

1. Shu Y, McCauley J. GISAID: Global initiative on sharing all influenza data – from vision to reality. Eurosurveillance 2017;22:30494.

2. Wickham H. ggplot2. Wiley Interdiscip Rev Comput Stat 2011;3:180–185.

